# Whole genome sequencing as an investigational device for return of hereditary disease risk and pharmacogenomic results as part of the *All of Us* Research Program

**DOI:** 10.1101/2021.04.18.21255364

**Authors:** E Venner, D Muzny, JD Smith, K Walker, CL Neben, CM Lockwood, PE Empey, GA Metcalf, The All of Us Research Program Regulatory Working Group, S Mian, A Musick, H Rehm, S Harrison, S Gabriel, R Gibbs, D Nickerson, AY Zhou, K Doheny, B Ozenberger, SE Topper, NJ Lennon

**Author notes:** These authors contributed equally (E Venner, D Muzny).

## Abstract

The *All of Us* Research Program (AoURP, ‘the program’) is an initiative, sponsored by the National Institutes of Health (NIH), that aims to enroll one million people (or more) across the United States. Through repeated engagement of participants, a research resource is being created to enable a variety of future observational and interventional studies. The program has also committed to genomic data generation and returning important health-related information to participants. To do so, whole genome sequencing (WGS), variant calling processes, data interpretation, and return-of-results procedures had to be created and receive an Investigational Device Exemption (IDE) from the United States Food and Drug Administration (FDA). The performance of the entire workflow was assessed through the largest known cross-center, WGS-based, validation activity that was refined iteratively through interactions with the FDA over many months. The accuracy and precision of the WGS process as a device for the return of certain health-related genomic results was determined to be sufficient, and an IDE was granted. We present here both the process of navigating the IDE application process with the FDA and the results of the validation study as a guide to future projects which may need to follow a similar path. Future supplements to the IDE will be submitted to support additional variant classes, sample types, and any expansion to the reportable regions.

## Background

The primary objectives of the *All of Us* Research Program (AoURP, ‘the program’) are: 1) to build a comprehensive research resource composed of surveys, biometrics, genetics, electronic health records, and biospecimens from one million or more participants reflecting the diversity of the United States; 2) to make these data and biospecimens broadly available for research exploring biological, social, and environmental determinants of health and disease; and 3) to return genomics results, gleaned from whole genome sequencing (WGS) (1) or genotyping arrays, directly to participants who elect to receive such information. Although some previous research studies have returned genomics results to their participants (2–9) and many clinical laboratories have validated WGS pipelines for the purpose of returning germline disease diagnoses (10), none have done so on the scale, or with the diversity of participants, as the AoURP.

The program adopted the genes identified by the American College of Medical Genetics and Genomics (ACMG) for return of incidental findings (11) as the returnable regions for the ‘Hereditary Disease Risk (HDR) Report’. Additionally, portions of seven genes with known gene-drug interactions were chosen for return in the ‘Medicine and Your DNA Report’ (hereafter referred to as the Pharmacogenomics or ‘PGx Report’). An overview of the genomics workflow for return of results is shown in Figure 1.

**Figure 1.**
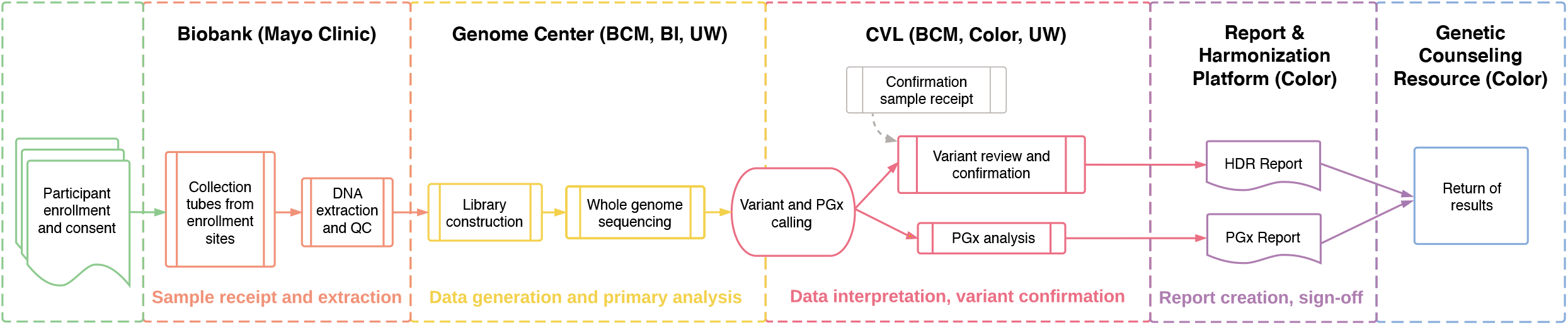
Overview of *All of Us* Research Program return of genomic results workflow. Participants are enrolled at a variety of locations including mobile sites, hospitals, and walk-in clinics. Samples are sent to the Biobank (Mayo Clinic) where DNA is extracted and stored. Upon indication from the program to proceed, samples are sent to one of three Genome Centers – Baylor College of Medicine (BCM), Broad Institute (BI), or University of Washington (UW) – for whole genome sequencing. For participants who have consented to return of genomic results, data are forwarded on to the Clinical Validation Laboratories (CVLs) for pharmacogenomics (PGx) analysis and variant interpretation and orthogonal confirmation for Hereditary Disease Risk (HDR) gene pathogenic or likely pathogenic variants. Reports are generated by the Report and Harmonization Platform (Color) and delivered to participants through the Genetic Counseling Resource (Color). QC, quality control.

During the conceptualization of the AoURP, NIH staff consulted with staff at the United States Food and Drug Administration (FDA), who determined that the proposed project met criteria for a Significant Risk (SR) Device Study, defined as incorporating a device that “is for a use of substantial importance in diagnosing, curing, mitigating, or treating disease, or otherwise preventing impairment of human health and presents a potential for serious risk to the health, safety, or welfare of a subject” (21 CFR 812.3). Because of this, an Investigational Device Exemption (IDE) would be required for the return of health-related genomic results in addition to institutional review board (IRB) approval (12). The requirement for FDA approval of NIH-sponsored research projects intending to return genetic results has been the topic of much prior discussion, primarily due to concerns over jurisdiction (i.e., FDA vs. IRB) and the non-trivial process of defining the exact requirements for approval (13). Previous FDA approvals of direct-to-consumer genomic sequencing tests (such as Foundation Medicine (14) and 23andMe (14)) and return of results from research projects (such as NSIGHT (13)) served as useful examples, but the AoURP presented a unique set of challenges and, with its high visibility, an opportunity to establish precedents for the genomic medicine research community.

Here we present the IDE process, the results of the analytical validity analyses performed to satisfy the IDE requirements, and the lessons learned to benefit future studies seeking to follow a similar path. Additional analyses that were performed, but outside the scope of this work, include gene and variant selection criteria, evidence justifications for PGx gene-drug associations, methods for variant harmonization, and comprehension testing of the HDR and PGx Reports.

## Methods

### Interrogated regions of the genome

The portion of the whole genome that was proposed for the return of genomic results encompassed 223,913 bases across 66 genes. Of these, 59 and 7 genes comprise the HDR, and PGx reports, respectively (Table S1).

### Determination of validation requirements and validation samples used

Requirements were refined in collaboration with the FDA. We identified 1,299 DNA samples, both whole blood and buffy coat and human-derived cell lines (Table 1). 217 samples were shareable across the AoURP Genome Centers (Baylor College of Medicine, BCM; Broad Institute, Broad; and University of Washington, UW) and therefore repeated at each center, and 350 center-specific patient samples were run only at a single site. The shared samples included a set of standard cell line controls (e.g., NA12878, Ashkenazi trio), cell line samples selected for specific PGx and ACMG59 variants (Coriell Biorepository), and commercially sourced, donor blood samples housed at the AoURP Biobank (Mayo Clinic) to validate extraction methods. The center-specific clinical samples were primarily used to determine accuracy for detection of specific ACMG59 and PGx alleles. In addition to clinical samples, we assessed accuracy on PGx alleles using DNA from the Genetic Testing Reference Materials Coordination Program (GeT-RM) (15), which provides extensively characterized PGx cell lines. For reportable PGx alleles that were not present in the GeT-RM collection or in clinical samples, we procured and processed cell lines that were part of the 1000 Genomes Project and had had PGx calling done as part of that effort.

**Table 1.** Summary of assessments and numbers of specimens included in each study

| Sample cohort | Assessment |  |  |  |  |  |  |  |
| --- | --- | --- | --- | --- | --- | --- | --- | --- |
|  | Reportable range | Accuracy | Precision | Interlab concordance | Limit of detection | Extraction performance | Molecular index performance | Assay fail rate |
| Blood-derived patient samples | n/a | 430 | n/a | n/a | n/a | n/a | n/a | 30 |
| Blood-derived healthy donor aliquots | 5 | n/a | 28 | 20 | 21 | 15 | 28 | 28 |
| GIAB<sup>a</sup> reference cell lines | 1 | 1 | n/a | n/a | 37 | n/a | 37 | 37 |
| Cell lines with known HDR<sup>b</sup> variants | 30 | 30 | 30 | 30 | n/a | n/a | 30 | 30 |
| CDC<sup>c</sup> GeT-RM<sup>d</sup> PGx<sup>e</sup> cell lines | 68 | 164 | 62 | 175 | n/a | n/a | 164 | 175 |
| Historical blood-derived WGS<sup>f</sup> data | n/a | n/a | n/a | n/a | n/a | n/a | n/a | 25,028 |

<sup>a</sup>GIAB, Genome in a Bottle. <sup>b</sup>HDR, Hereditary Disease Risk. <sup>c</sup>CDC, United States Centers for Disease Control and Prevention. <sup>d</sup>GeT-RM, Genetic Testing Reference Materials Coordination Program. <sup>e</sup>PGx, pharmacogenomics. <sup>f</sup>WGS, whole genome sequencing.

### Sample preparation, sequencing, and primary and secondary bioinformatics

For the control samples, we received DNA from Coriell Biorepository. DNA from commercially-sourced blood samples and future AoURP participant samples was extracted from 4 ml EDTA (whole blood) or 10 ml EDTA (buffy coat) by two methods: salt-based precipitation on Autogen FlexStar or bead-based method on Chemagen 360. Samples were stored in −80°C automated freezers and were checked for volume via a BioMicroLab volume check instrument. DNA samples were also quantified (spectrometric method) via Lunatic-Unchained Labs / Trinean DropSense 96 to obtain total DNA concentration as well as A260/280 and A260/230. All samples met a minimum concentration of 50 ng/ul and an A260/280 of 1.6-2.0. To assess performance equivalency of the two methods, whole blood and buffy coat (WBC) specimens from five donors were extracted using autogen and chemagen platforms. DNA from these samples was sequenced using WGS and an orthogonal targeted panel assay.

Each Genome Center performed quality control (confirmation of volume and concentration) of the samples submitted from the AoURP Biobank. Samples that met quality thresholds were accessionned and sample aliquots were prepared for library construction processing (normalized with respect to concentration and volume).

DNA samples were first sheared using a Covaris sonicator and then size-selected using AMPure XP beads to restrict the range of library insert sizes. Libraries were constructed using the PCR Free Kapa HyperPrep library construction kit and utilizing dual-indexed adapters. Libraries were quantified using qPCR with the Illumina Kapa DNA Quantification Kit and then normalized and pooled for sequencing. Actual implementations of the library construction processes (automation platforms used, for instance) varied across the Genome Centers. Pooled libraries were loaded on the Illumina NovaSeq 6000 instrument, and WGS was performed with Illumina reagents following the manufacturer’s best practices.

After demultiplexing, WGS analysis occurred on the DRAGEN Platform (Illumina), which consists of optimized algorithms for mapping, aligning, sorting, duplicate marking, and haplotype variant calling. Alignment used the GRCh38DH reference genome. The DRAGEN pipeline produced a large number of metrics that cover lane, library, flow cell, barcode, and sample-level metrics for all runs as well as assessing contamination and mapping quality. For the purposes of the IDE analyses, the software version of the DRAGEN software was harmonized to the 3.4.12 version at all Genome Centers.

### Data analyses

#### Accuracy

In the absence of an FDA-approved ground truth assay for each reportable variant, the FDA requested that accuracy be presented as the positive and negative percent agreement (PPA and NPA, respectively) of the device calls compared to a high quality comparator assay. For comparator assays, we used clinically validated gene panels and in some cases capillary sequencing. In the case where the gene panel data was used as a comparator, calls made from the panel were designated as ‘true positive’ and ‘true negative’, and calls from the WGS were categorized based on presence or absence in the panel data. Specific important variants in the population were represented in the dataset, including founder alleles in *BRCA1* and *BRCA2* (Table S2).

#### Calculation of PPA and NPA in clinical specimens

A set of 271 patient samples were examined for accuracy across a range of genomic contexts, variant subtypes, and zygosity. Gene panels were those used in the eMERGE III study at BCM and Broad (16) or an ACMG panel at UW (17), intersected with the HDR Report interval. Genomic contexts were defined using bed files from the Genome Alliance for Genomic Health (GA4GH, www.ga4gh.org) benchmarking-tools repository.

#### Concordance of pathogenic/likely pathogenic calls in cell lines

Previously characterized human cell line-derived DNA (Coriell Biorepository) was processed through the production workflows for WGS, capillary, or panel sequencing at all Genome Centers and CVLs (BCM; UW; and Color Health, Color). Calls of the known pathogenic (P) or likely pathogenic (LP) variants were assessed at each site and concordance measured (Table S3).

#### Performance in reference cell lines

Performance on the National Institute of Standards and Technology (NIST) human reference cell line (NA12878) was evaluated by comparing to the Genome in a Bottle v3.3.2 gold standard truth set.

#### Accuracy of PGx sites

Accuracy of PGx calling was determined using 159 patient samples that had previously been orthogonally validated by Sanger sequencing, next generation sequencing (NGS) panel, or genotyping assays (Table 3). As noted above for the HDR genes, the selected PGx clinical samples provided a representation of genes and alleles that aligned with the expected prevalence of reportable alles in the general population as determined by Clinical Pharmacogenetics Implementation Consortium (CPIC) (www.cpicpgx.org). In genes where reported allele frequencies are extremely rare and no clinical samples were accessible, cell line samples were used to provide a more comprehensive list of reportable alleles for validation (Table S4 and Table S5).

**Table 3.** Call concordance of PGx alleles using WGS and orthogonal methods in patient samples

| Gene and star allele | clinical samples | Correct calls | Incorrect calls | Correct calls | Incorrect calls | Correct calls | Incorrect calls | Correct calls | Incorrect calls | Correct calls | Incorrect calls | Correct calls | Incorrect calls | Correct calls | Incorrect calls | concordance |
| --- | --- | --- | --- | --- | --- | --- | --- | --- | --- | --- | --- | --- | --- | --- | --- | --- |
| CYP2C19*10 | 1 | 1/1 | 0 | - | - | - | - | 1/1 | 0 | - | - | - | - | - | - | 2/2 |
| CYP2C19*17 | 40 | 25/25 | 0 | 15/15 | 0 | - | - | 25/25 | 0 | 15/15 | 0 | - | - | - | - | 80/80 |
| CYP2C19*17/*17 | 1 | 1/1 | - | - | - | - | - | 1/1 | - | - | - | - | - | - | - | 2/2 |
| CYP2C19*2 | 48 | 33/33 | 0 | - | - | 15/15 | 0 | 33/33 | - | - | - | - | - | 15/15 | 0 | 96/96 |
| CYP2C19*2/*2 | 3 | 3/3 | 0 | - | - | - | - | 3/3 | - | - | - | - | - | - | - | 3/3 |
| CYP2C19*22 | 1 | - | - | 1/1 | 0 | - | - | - | - | 1/1 | 0 | - | - | - | - | 2/2 |
| CYP2C19*3 | 3 | 3/3 | 0 | - | - | - | - | 3/3 | - | - | - | - | - | - | - | 3/3 |
| CYP2C19*35 | 1 | 1/1 | 0 | - | - | - | - | 1/1 | - | - | - | - | - | - | - | 1/1 |
| CYP2C19*4 | 6 | 3/3 | 0 | 3/3 | 0 | - | - | 3/3 | 0 | 3/3 | 0 | - | - | - | - | 12/12 |
| CYP2C19*8 | 1 | - | - | - | - | 1/1 | 0 | - | - | - | - | - | - | 1/1 | 0 | 2/2 |
| CYP2C19*9 | 2 | - | - | 2/2 | 0 | - | - | - | - | 2/2 | 0 | - | - | - | - | 4/4 |
| DPYD c.1129-5923C>G | 2 | 2/2 | 0 |  |  |  |  | 2/2 | - | - | - | - | - | - | - | 4/4 |
| DPYD c.1679T>G (*13) | 3 | 3/3 | 0 |  |  |  |  | 3/3 | - | - | - | - | - | - | - | 6/6 |
| DPYD c.2846A>T | 2 | - | - | 2/2 | 0 | - | - | - | - | 2/2 | 0 | - | - | - | - | 4/4 |
| DPYD*2 (c.1905+1G>A) | 2 | - | - | 2/2 | 0 | - | - | - | - | 2/2 | 0 | - | - | - | - | 4/4 |
| G6PD A-202A_376G | 3 | 3/3 | 0 | - | - | - | - | 3/3 | 0 | - | - | - | - | - | - | 6/6 |
| G6PD Canton, Taiwan-Hakka, Gifu-like, Agrigento-like | 1 | 1/1 | 0 | - | - | - | - | 1/1 | 0 | - | - | - | - | - | - | 2/2 |
| G6PD<br>Ilesha | 2 | 2/2 | 0 | - | - | - | - | 2/2 | 0 | - | - | - | - | - | - | 4/4 |
| G6PD<br>Mediterranean, Dallas,<br>Panama,<br>Sassari,<br>Cagliari,<br>Birmingham | 5 | 5/5 | 0 | - | - | - | - | 5/5 | 0 | - | - | - | - | - | - | 10/10 |
| G6PD<br>Seattle,<br>Lodi,<br>Modena,<br>Ferrara II,<br>Athens-like | 1 | 1/1 | 0 | - | - | - | - | 1/1 | 0 | - | - | - | - | - | - | 2/2 |
| G6PD<br>Union,<br>Maewo,<br>Chinese-2,<br>Kalo | 2 | 2/2 | 0 | - | - | - | - | 2/2 | 0 | - | - | - | - | - | - | 4/4 |
| NUDT15*2 | 2 | 2/2 | 0 | - | - | - | - | 2/2 | 0 | - | - | - | - | - | - | 4/4 |
| NUDT15*3 | 9 | 8/8 | 0 | - | - | 1/1 | 0 | 8/8 | 0 | - | - | - | - | 1/1 | 0 | 18/18 |
| SLCO1B1*<br>15 | 28 | 22/22 | 0 | - | - | 6/6 | 0 | 22/22 | 0 | - | - | - | - | 6/6 | 0 | 56/56 |
| SLCO1B1*<br>15/*15 | 4 | 4/4 | 0 | - | - |  |  | 4/4 | 0 | - | - | - | - | - | - | 8/8 |
| SLCO1B1*<br>17 | 3 | - | - | - | - | 3/3 | 0 | - | - | - | - | - | - | 3/3 | 0 | 6/6 |
| SLCO1B1*<br>5 | 8 | 7/7 | 0 | - | - | 1/1 | 0 | 7/7 | 0 | - | - | - | - | 1/1 | 0 | 16/16 |
| TPMT*2 | 6 | 5/5 | 0 | 1/1 | 0 | - | - | 5/5 | 0 | 1/1 | 0 | - | - | - | - | 12/12 |
| TPMT*3A | 9 | 9/9 | 0 | - | - | - | - | 9/9 | 0 | - | - | - | - | - | - | 18/18 |
| TPMT*3C | 5 | 5/5 | 0 | - | - | - | - | 5/5 | 0 | - | - | - | - | - | - | 10/10 |
| UGT1A1*2<br>7 | 3 | 3/3 | 0 | - | - | - | - | 3/3 | 0 | - | - | - | - | - | - | 6/6 |
| UGT1A1*2<br>8 | 70 | 37/37 | 0 | 33/33 | 0 | - | - | 37/37 | 0 | 33/33 | 0 | - | - | - | - | 140/140 |
| UGT1A1*3<br>6 | 12 | 4/4 | 0 | 8/8 | 0 | - | - | 4/4 | 0 | 8/8 | 0 | - | - | - | - | 24/24 |
| UGT1A1*3<br>6/*36 | 1 | 1/1 | 0 | - | - | - | - | 1/1 | 0 | - | - | - | - | - | - | 2/2 |
| UGT1A1*3<br>7 | 3 | 2/2 | 0 | - | - | 1/1 | 0 | 2/2 | 0 | - | - | 1/1 | 0 | - | - | 6/6 |
| UGT1A1*6 | 8 | 3/3 | 0 | 5/5 | 0 | - | - | 3/3 | 0 | 5/5 | 0 | - | - | - | - | 16/16 |
|  |  | Broad<sup>a</sup><br>(WGS<sup>b</sup>) |  | BCM<sup>c</sup> (WGS) |  | UW<sup>d</sup> (WGS) |  | Color<sup>e</sup> CVL<sup>f</sup><br>(Panel) |  | BCM CVL<br>(Sanger) |  | UW CVL<br>(Panel) |  | UW CVL<br>(Array) |  |  |

<sup>a</sup>BI, Broad Institute. <sup>b</sup>WGS, whole genome sequencing. <sup>c</sup>BCM, Baylor College of Medicine. <sup>d</sup>UW, University of Washington. <sup>e</sup>Color, Color Health. <sup>f</sup>CVL, Clinical Validation Laboratory.

#### Precision

To assess equivalence of processing and variant calling across the three Genome Centers within the AoURP device, we computed the concordance of variant calls from five donor-derived blood specimens collected and processed through the WGS and variant calling pipelines. Replicate samples were run at each center, and the equivalence between replicates was determined to demonstrate that the variability in sample processing and variant calling between labs was no greater than the variability within labs (Table S6). Overall equivalence was calculated using the Jaccard similarity coefficient between each pair of labs over all variants (i.e., the size of the intersection of the calls divided by the size of the union of the calls). We further evaluated equivalence across and between labs using the WGS data from 175 human cell line derived genomic DNA samples that were part of the PGx accuracy and NIST accuracy studies (Table S7).

#### Inter- and intra-Genome Center precision

Two studies were completed to calculate inter- and intra-Genome Center precision. In the first study, 20 replicate blood-derived DNA samples from five individual donors were examined. Clinical gene panel testing (18,19) of these samples was used to define the ‘truth’. Variant calls from WGS on all 20 samples at each Genome Center were compared to the panel variants to determine concordance across sites by genomic context (Table S8). In the second study, 30 human cell lines (the same cell lines as were used in the P/LP variant accuracy assessment) were sequenced utilizing the clinical NGS panel as described above to define the ‘truth.’ Calls from WGS on all 30 samples at each Genome Center were compared to the panel variants to determine concordance across sites by genomic context (Table S9). To demonstrate the equivalence of cell line-derived DNA with that of blood-derived primary samples, we calculated performance measures and technical measures for selected assessments, run on both clinical cohorts and cell lines (Table S10 and Table S11).

#### Precision of PGx calling

Precision of PGx variant calling was assessed by processing 62 cell lines with known PGx alleles, as defined by Stargazer (20) at each of the three Genome Centers and CVLs (Table S12).

#### Limit of detection

In order to determine the range of acceptable genomic DNA inputs into library construction, an input titration experiment using DNA derived from NA12878 with total input amounts ranging from 25 ng to 1500 ng into library construction was performed. These input levels span a range from 10X the lowest acceptable input to 2X the highest standard input into library construction across the Genome Centers. To assess the effect of lower input amounts on sensitivity and to confirm that the minimum input identified produces acceptable sensitivity and precision, the vcfeval tool (21) was leveraged to calculate sensitivity versus NIST for each titration point (Figure S6). A similar titration and analysis was done with samples from the AoURP Biobank (Table S13).

#### Sequence generation quality control specification determination

For coverage metrics, we used the Picard tool to remove read data uniformly (downsampling) from four NIST control samples for which gold-standard variant data is available. For contamination, we created a set of bioinformatically contaminated samples by adding progressively more read data from a second sample to a control sample. For the duplicate rate we progressively added more duplicates to a sample while holding the yield constant.

#### Reportable range

To define reportable genomic intervals we selected transcripts for each of the 59 genes of interest, using primarily MANE Select (22) and RefSeq (23) guidelines. We extend exons from the −15 upstream intronic position to the +6 downstream intronic position. We then added intervals to cover known P/LP variants outside of the −15 to +6 regions and for 43 PGx star allele sites. We excluded three types of technically challenging regions from the reportable range: 1) *PMS2* exons 12-15, which has high homology with the *PMS2CL* pseudogene; 2) regions with high GC content (typically >75% across 100 bp) that can result in low coverage; and 3) regions with spurious variant calling artifacts due to the presence of micro-repeats (di-, trinucleotides) and long homopolymers.

An additional set of regions did not consistently have at least 20X coverage in 20% of the samples in the dataset. A per-site coverage analysis was performed across the entire HDR gene and PGx site region with a dataset of 104 samples from the three Genome Centers with a whole-genome mean coverage of 30-35X. This analysis revealed six regions that included 56 bases across four genes (Table S14). For these regions, none were low enough quality to consider excluding the region from the reportable range.

#### Invalid rates

To illustrate fail rates at each step, we used historical data and data from this validation study Invalid rates were also calculated from panel data at Color and UW. BCM data represents capillary sequence data from an internal cohort. For historical data, we used the National Heart, Lung, and Blood Institute Trans-Omics for Precision Medicine (TOPMed) cohort as it represents a large number of samples run at all three Genome Centers (Table S15 and Table S16).

## Results

### Initial assessment of FDA requirements

Over 19 months, the Genome Centers, CVLs and NIH staff discussed each major element of the program (including participant consent, sample processing, analysis and interpretation, and return of results) with reviewers at the FDA via a series of conference calls and presubmission inquiries, to define the elements required for the IDE application (Figure 2). The FDA determined that the ‘device’ to be approved in this case constituted all steps of the process and requested that validation samples be primarily derived from blood to match the sample types used by the program. Those samples should reflect the variant types (single nucleotide variants, SNVs, and insertions or deletions, indels) and genomic properties (GC rich, complexity) that were anticipated to be reported to participants. After submission of the IDE, the FDA requested clarifications and analyses that are summarized in Table S17. An analysis of the incidence of reportable variants within the HDR genes across the clinical laboratories at BCM, Color, UW, and Laboratory for Molecular Medicine at Partners HealthCare (LMM, associated with the Broad Genome Center) revealed that some genes will likely have very few reportable variants and that indels are not reportable in several genes (Figure S1).

**Figure 2.**
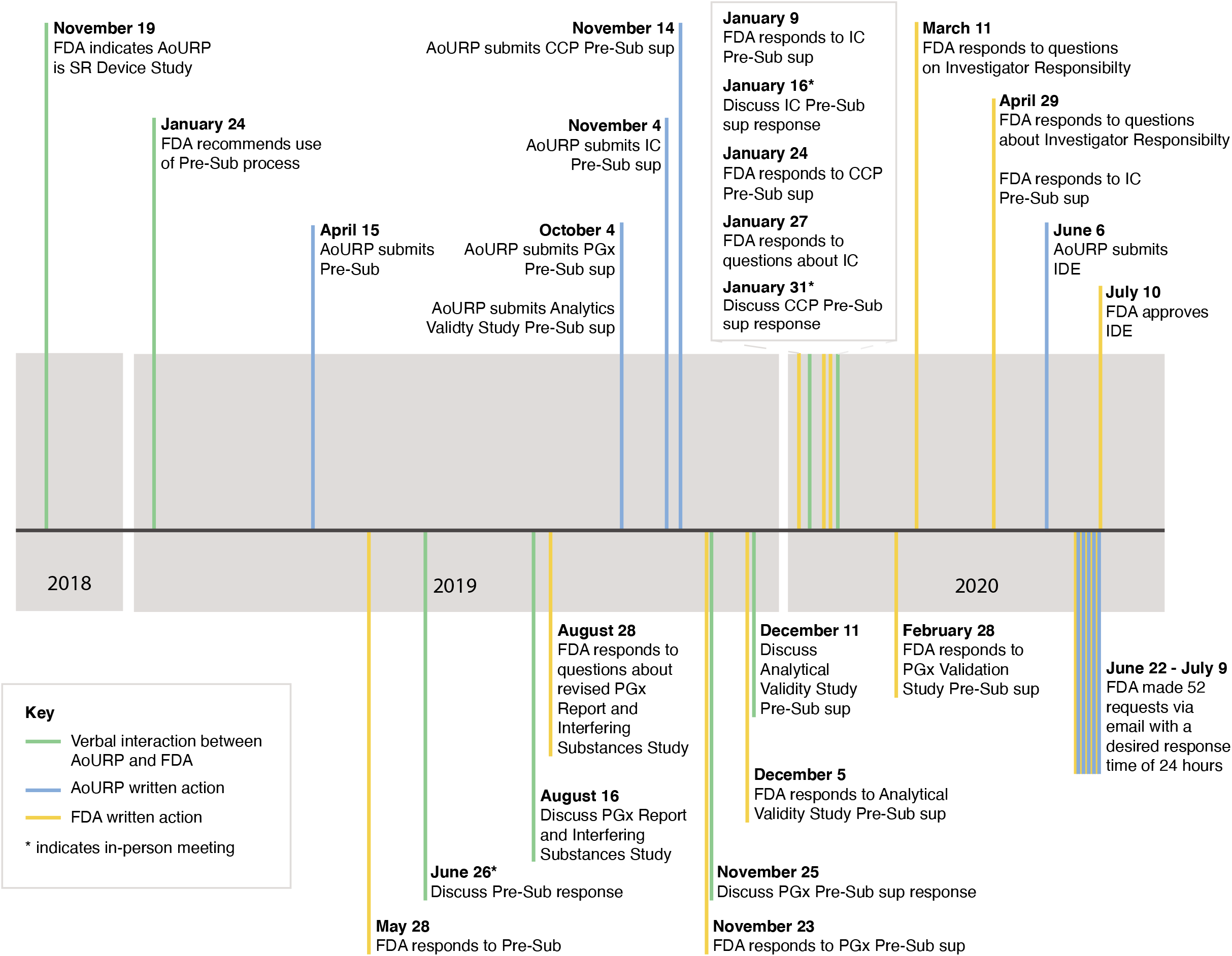
Timeline of the IDE application process. The requirements for the investigational device exemption (IDE) content were refined through a series of pre-IDE submissions and responses, in-person meetings, and teleconferences with the United States Food and Drug Administration (FDA) over a period of 19 months. AoURP, *All of Us* Research Program. CCP, Change Control Policy. IC, informed consent. PGx, pharmacogenomics. Pre-Sub, pre-submission. SR, significant risk. Sup, supplement.

Per the IDE process, requirements were set by the study sponsor (NIH), which were then used to define five specific acceptance criteria. The data showed sequencing depth exceeding 20X for the HDR and PGx reportable range, high accuracy for all Genome Centers (>99.7%), and high concordance of data generated at the Genome Centers (>99.7%) for both the HDR region and PGx alleles. Limit-of-detection studies demonstrated accurate data produced from a range of input amounts with an expected invalid rate for blood-derived specimens of <1%.

### Accuracy

Across a set of 271 clinical samples, PPA ranged from 94.24% to 100% when broken down by genomic context, with NPA at 100% across all categories (Table 2 and Table S8). The accuracy of the AoURP device was >99% (horizontal dotted line) in events up to 20 bases in length (Figure S2). We detected five recurrent false positive and false negative variants in a large number of the samples (Table S18) which were removed from the analysis. All were benign, likely benign or variants of uncertain significance, would not be reported clinically, and fall in regions that are known to have sequence homology or mapping issues (24). On a set of human cell line controls, overall concordance was 100% for a set of known P/LP variants (Table S3), and in a detailed assessment of the NA12878 control sample variant calling, accuracy was 100% for variants in the reportable range (Table S19) and 99.89% for variants across the whole genome (Table S20). Accuracy of PGx calling was determined on 159 blood-derived patient samples which had previously been orthogonally validated in each Genome Center (Table 3). We observed concordance in 100% of calls (595/595).

**Table 2.** Accuracy of variant calling across genomic contexts and variant types in patient samples

|  | **Panel+<sup>a</sup>/**<br>**WGS–<sup>b</sup>** | **Panel–/**<br>**WGS+** | **Panel+/**<br>**WGS+** | **Panel–/**<br>**WGS–** | **PPA<sup>c</sup> [95% CI<sup>d</sup>]** | **NPA<sup>e</sup> [95% CI]** |
| --- | --- | --- | --- | --- | --- | --- |
| All compiled | 76<sup>h</sup> | 201 | 29,475 | 45847148 | 99.74%<br>[99.68%-99.81%] | 100%<br>[100%-100%] |
| P/LP<sup>f</sup> variants only | 0 | 0 | 145 | 28474503 | 100%<br>[100%-100%] | 100%<br>[100%-100%] |
| SNVs<sup>g</sup> only | 23 | 6 | 28,710 | 45848108 | 99.92%<br>[99.87%-99.97%] | 100%<br>[100%-100%] |
| Insertions only | 9 | 8 | 225 | 29887204 | 96.20%<br>[94.30%-98.00%] | 100%<br>[100%-100%] |
| Deletions only | 33 | 12 | 540 | 42170158 | 94.24%<br>[92.65%-95.83%] | 100%<br>[100%-100%] |
| Segmental duplications | 12 | 2 | 2,119 | 38466267 | 99.44%<br>[99.10%-99.78%] | 100%<br>[100%-100%] |
| Known pseudogenes | 9 | 74 | 1,075 | 45676596 | 99.17%<br>[98.60%-99.73%] | 100%<br>[100%-100%] |
| Low mappability regions | 18 | 12 | 1,069 | 36912107 | 98.34%<br>[97.84%-98.85%] | 100%<br>[100%-100%] |
| Low complexity regions | 24 | 69 | 956 | 38072735 | 97.55%<br>[96.35%-98.75%] | 100%<br>[100%-100%] |
| Low GC regions | 6 | 12 | 197 | 14792575 | 97.04%<br>[95.87%-98.22%] | 100%<br>[100%-100%] |
| High GC regions | 0 | 0 | 23 | 3891553 | 100%<br>[100%-100%] | 100%<br>[100%-100%] |
| Heterozygous variants only | 75 | 147 | 18,295 | 45858382 | 99.59%<br>[98.48%-99.70%] | 100%<br>[100%-100%] |
| Homozygous variants only | 1 | 83 | 11,180 | 45865561 | 99.99%<br>[99.97%-100%] | 100%<br>[100%-100%] |

<sup>a</sup>+ indicates that a variant was present on the panel or the corresponding whole genome sequencing (WGS) sample. <sup>b</sup>– indicates that a variant was not present on the panel or the corresponding WGS sample. <sup>c</sup>PPA, positive percent agreement. <sup>d</sup>CI, confidence
interval. <sup>e</sup>NPA, negative percent agreement. <sup>f</sup>P/LP, pathogenic/likely pathogenic. <sup>g</sup>SNVs, single nucleotide variants. <sup>h</sup>Subsequent to the investigational device exemption (IDE) submission, additional review indicated that this number should in fact be a total 21 panel positive, but WGS negative variants distributed across the various subcategories (some variants were missannotated in the original tables). However, to reflect what was actually submitted as part of the IDE, the original number is listed here. The impact on overall performance measures is small.

### Precision

The Genome Centers jointly agreed to harmonize the production pipelines to the extent possible, but some parts of the pipelines (e.g., DNA quality control [QC] and quantification) could not be harmonized perfectly, so we used sample replicates to demonstrate functional equivalence. 20 blood samples from five individuals were examined, with clinical, gene-panel testing of these samples used to define ‘truth’. WGS variants were compared to the panel variants to determine concordance across sites and discordance by genomic context (Table S8). The data were highly concordant, with the differences observed between Genome Centers smaller than the within-center sample variance. When broken out by genomic context, the context with the largest range was insertions, where PPA varied between 94.44% and 100%.

PGx variant calling precision across the three Genome Centers was assessed by processing 62 cell lines with known PGx alleles. A total of 298 of 300 alleles were found to be concordant, with discordance only found in *G6PD* alleles due to an incorrect ploidy call in the bioinformatics pipeline (Table S12).

### Limit of detection

The minimum amount of input DNA for sequencing was determined from NA12878 to be 250 ng, as lower amounts either failed to produce a library or showed reduced sensitivity. Additionally, four donor blood samples from the AoURP Biobank were compared against that sample’s gene panel data, with assessments of sensitivity and precision (Table S13). Across a range of extraction methods, we observed a range of PPA from 98.1% for autogen indels to 100% for chemogen indels. NPA remained at 100% across these comparisons.

To evaluate performance as a function of allele fraction, seven replicates of NA12878 were sequenced with the AoURP WGS pipeline and compared to results from NIST, restricted to the high confidence regions. The analysis indicates that confident and accurate heterozygous calls are made between 30-75% allele fraction for SNVs and 30-65% allele fraction for indels (Figure S8).

### Other Analyses

We performed several additional analyses to satisfy the FDA requirements. We assessed equivalent performance and technical measures of cell line-derived DNA with that of blood-derived DNA. Clinical specimens and cell lines were highly concordant (Table S10 and Table S11). We demonstrated that the intended analyte was being measured by noting that the K_2_EDTA blood collection kits used by the program were approved by the FDA for hematological blood analysis. We described how quality-metric thresholds were selected and optimized (Figures S3-S4), showed that variant performance from both extraction methods and both blood collections were equivalent (Table S21), and determined a fail rate below 1% for blood-derived specimens. Finally, we established the accuracy of the ‘liftover’ step (conversion of variants called on the GRCh38DH reference to the GRCh37 reference), with the exception of four sites that had no corresponding GRCh37 position (Table S22).

### Change management

In concordance with FDA regulations (25), the AoURP established a Change Control Policy, which stated that each proposed change will be assessed for risk by an expert review panel made up of members from the AoURP Genome Centers and CVLs, and a formal recommendation is submitted for NIH evaluation and approval prior to FDA involvement. Categories are summarized in Table S23. For major changes (e.g., new reportable genes, changing procedure QC metrics and acceptance criteria, addition of new concepts to reports), the AoURP will obtain FDA approval through a supplemental application to the parent IDE. ‘Moderate’ changes, defined as those that do not affect the validity of the data (see 21 CFR 812.35(a)(3)(ii) for complete definition) require FDA notification within 5 working days of implementation. Minor changes (defined fully in 21 CFR 812.150(b)(5)) may be reported to the FDA in the annual report.

## Discussion

The FDA IDE process for the return of genomic results required nearly two years to complete but ultimately demonstrated that WGS as a clinical laboratory assay performed at a high level across all three Genome Centers, genes of interest, and various sequence contexts. Based on previous FDA submissions from Foundation Medicine(26), MSK-IMPACT(27), and 23andMe(28), as well as the experience in the NSIGHT project (13) and the FDA’s guidelines (29), we initially expected the requirements for the IDE process to be largely similar to the Clinical Laboratory Improvement Amendments (CLIA) validation framework under which the AoURP Genome Centers and CVLs operate their clinical laboratories (Table S24). Instead, we found that the FDA requirements exceed those that are required by CLIA.

Genomic testing research studies under the supervision of IRB protocols, as the AoURP is, have historically not been subject to IDE regulation (30). However, 2013 marked a distinction from this position when the FDA asserted authority over the use of genomic sequence of newborns with the NSIGHT project (13). Our experience has shown that any group initiating a research project that may require FDA approval should budget considerable upfront time and resources to the process. This is particularly true if the ‘device’ has no predicate in FDA approval history. The complexity of defining, designing, and executing on a validation study of this scale ultimately required that we reduce the proposed scope in several important ways (e.g., limiting acceptable specimen input types to blood, returning only SNVs and Indels, reducing reportable PGx alleles).

A major impediment to efficient approval was the lack of a closely related predicate test (i.e., a test that has previously been reviewed and approved by the FDA). Previously reviewed NGS and genotyping assays (e.g., Foundation Medicine, 23andMe) were different enough as to not be considered predicates. The FDA staff extrapolated from tests and technologies that they had previously reviewed, such as genotyping arrays and targeted sequencing.

Professional organizations provide guidelines for clinical laboratories who are validating genomic assays (31)(32). The FDA’s approach differs from these guidelines in important ways. First, the definition of ‘ground truth’ that the FDA strongly preferred required patient-derived clinical specimens over reference samples. Second, assessing variant calling performance for a particular variant class in a representative number of sites is generally considered acceptable as a proxy for performance of that variant class at other genomic positions (31)(32). However, the FDA required us to demonstrate performance in every reportable gene and PGx allele. The three Genome Centers collectively had access to hundreds of clinical remnant specimens from previous studies; for a smaller group or a single laboratory applying for an IDE, this would have been a challenging requirement.

This work represents the first time a sequencing consortium has harmonized the bioinformatics pipelines at the level of the software version and command-line parameters instead of focusing on ‘functional equivalence’. This allowed us to capitalize on one another’s validations and greatly simplified concordance calculations. However, one potential downside is the lack of ‘multiple views’ of the same data set, provided by multiple independent analysis approaches, which can support one another where they agree and potentially reveal systematic problems where they do not.

The AoURP is a groundbreaking research project that will generate a massive dataset to accelerate the study of disease but which also presents challenges under the current regulatory landscape. After a multi-year effort across multiple groups, the AoURP device was validated and an IDE was granted so that the genomics arm of the program could begin. We appreciate the collaborative relationship with the FDA and hope that this work will provide a streamlined model for future projects.

## Data Availability

Clinical sample data from this study are not publicly available.

## Acknowledgements

The AoU regulatory work group includes the following members: Anji Addington, Jill Alfodi, Beth Collins, Colleen Davis, Kim Doheny, Shannon Dugan-Perez, Chris Frazar, Chris Gocke, Namrata Gupta, Simon Hearsey, Hannah Hoban, Lawrence Hon, Elvin Hsu, Jameson Hurless, Viktoriya Korchina, Christie Kovar, Katie Larsson, Niall Lennon, Tina lockwood, Ginger Metcalf, Sana Mian, Mullai Murugan, Donna Muzny, Cynthia Neben, Chris O’Donnell, Anju Ondov, Brad Ozenberger, Aparna Pallavajjalla, Heidi Rehm, Jessica Reusch, Josh Smith, Monica Sulit, Scott Topper, Kimberly Walker, Anastasia Wise, Betty Woolf, and Alicia Zhou.

## Supplementary Tables

<u>Table S1. Reportable HDR and PGx genes</u>

<u>Table S2. Founder mutations in</u> *<u>BRCA1</u>*<u>and</u> *<u>BRCA2</u>*<u>called with 100% accuracy in independent samples</u>

<u>Table S3. Call concordance of select pathogenic variants in human cell lines</u>

<u>Table S4. Call concordance of PGx alleles using WGS and orthogonal methods in GeT-RM cell lines</u>

<u>Table S5. PGx calling across additional cell line samples from the 1000 Genomes Project</u>

<u>Table S6. Overall equivalence of called variants in donor blood samples across the Genome Centers</u>

<u>Table S7. Overall equivalence of called variants in cell lines across the Genome Centers</u>

<u>Table S8. Concordance of variant calling by genomic context in donor blood samples</u>

<u>Table S9. Concordance of variant calling by genomic context in cell lines</u>

<u>Table S10. Equivalence of performance measures between cell line-derived DNA and blood-derived DNA</u>

<u>Table S11. Equivalence of technical metrics between cell line-derived DNA and blood-derived DNA</u>

<u>Table S12. Concordance of PGx calling across the Genome Centers</u>

<u>Table S13. Input titration results for four blood donor samples</u>

<u>Table S14. Frequently underperforming bases within the Hereditary Disease Risk Report</u>

<u>Table S15. Fail rates for samples at discrete parts of the process across the Genome Centers</u>

<u>Table S16. Fail rates for samples at discrete parts of the process across the Clinical Validation Laboratories</u>

<u>Table S17. Feedback post IDE submission</u>

<u>Table S18. Excluded recurrent false positive and false negative variants</u>

<u>Table S19. Accuracy of variant calls across the reportable region of NA12878</u>

<u>Table S20. Accuracy of variant calls across the whole genome of NA12878</u>

<u>Table S21. Performance of different extraction platforms and input material types</u>

<u>Table S22. Mismatched bases after liftover between reference builds</u>

<u>Table S23. Change management process, by risk categorization</u>

<u>Table S24. Lessons learned during the pre-submission process</u>

## Supplemental Figures and Figure Legends

<u>Figure S1. Number of true positive variants observed in the Hereditary Disease Risk genes.</u> Data from a previous study was used to assess the likely frequency of reportable variants in the HDR genes. At least one true positive A) single nucleotide variant (SNV) or B) insertion or deletion (indel) was observed in each of the Hereditary Disease Risk (HDR) genes. Three additional genes not in the HDR gene list (*SLCO1B1, TMPT*, and *CYP2C19*) were included as they were on the orthogonal gene panels used for comparison. * indicates genes with potential surgical interventions indicated for a pathogenic/likely pathogenic variant. ✢ indicates genes in which loss of function is not a known mechanism of disease pathogenesis, meaning insertions or deletions are typically not reportable.

<u>Figure S2. Performance of variant calling from WGS as a function of insertion size and deletion size.</u> The accuracy of the AoURP device was >99% (horizontal dotted line) in events up to 20 bases in length (vertical dotted lines) and ≥97% in events out to 30 bases in length when compared to a well-established truth sample (NA12878). Red line indicates sensitivity, and blue line indicates precision.

<u>Figure S3. Sequence generation quality control specification determination.</u> We generated data to demonstrate thresholds used for QC metrics. The total A) false positives and B) false negatives showed a gradual increase as mean coverage decreased, with a rapid increase below 20X coverage. The total C) false positives and D) false negatives steadily increased as the percentage of bases with at least 20X coverage decreased. The total E) false positives and F) false negatives increased gradually as the average coverage in Hereditary Disease Risk Report (HDRR) regions decreased. The reduction in performance was slow initially and then increased rapidly below 40%. The total G) false positives and H) false negatives increased with lower base quality counts, with inflection points starting around 6e10 for both. Vertical purple line marks the device acceptance criteria.

<u>Figure S4. Relationship between estimated sample contamination and performance.</u> Variant calling performance, as measured by both the number of false positives (pos) and the number of false negatives (neg) for single nucleotide polymorphisms (SNPs) and insertions or deletions (indels), decreased with increasing contamination. Blue line indicates false positives, and red line indicates false negatives. Blue line indicates false positives, and red line indicates false negatives. Vertical purple line marks the device acceptance criteria.

<u>Figure S5. Relationship between duplicate rate and performance.</u> Samples with a higher percentage of duplicate reads had a higher total number of false positive (pos) and false negative (neg) single nucleotide polymorphisms (SNPs) and insertions or deletions (indels). Blue line indicates false positives, and red line indicates false negatives. Vertical purple line marks the device acceptance criteria.

<u>Figure S6. Relationship between mean coverage and uniformity.</u> The uniformity metric did not deviate as data quality was reduced in four well-established truth samples (NA12878, NA23143, NA24149, and NA24385).

<u>Figure S7. Analytical sensitivity for NA12878 input titration series vs NIST across all three Genome Centers.</u> Library input amounts below 250 ng showed reduced sensitivity. There was no significant difference in performance between 250 ng and 1500 ng input DNA. Red line indicates precision, and blue line indicates sensitivity. Indel, insertion or deletion. SNV, single nucleotide variant.

<u>Figure S8. Precision and recall (i.e., sensitivity) of SNV and indel calling as a function of alt allele fraction.</u> Heterozygous calls made between 30-75% allele fraction for single nucleotide variants (SNVs) and 30-65% allele fraction for insertions or deletions (indels) had high A) precision and B) recall. Peach line indicates indels, and teal line indicates SNPs. Alt, alternative.

## Notes

### Competing Interest Statement

EV is cofounder of Codified Genomics. All other authors declare no competing interests.

### Funding Statement

This work was funded by NIH awards 1OT2OD002751-01, 1OT2OD002750-01 and 1OT2OD002748-01.

### Author Declarations

AoU IRB approval v9: Mar 20, 2020 (https://allofus.nih.gov/about/who-we-are/institutional-review-board-irb-of-all-of-us-research-program)

