## Supplemental Table 1 for "Whole genome sequencing as an investigational device for return of hereditary disease risk and pharmacogenomic results as part of the *All of Us* Research Program"

Table S1. Reportable HDR and PGx genes

| **Gene** | **Drug (brand name) / Disease (MIM number)** | **Report type** |
| --- | --- | --- |
| *TPMT* | azathioprine (Imuran®)  mercaptopurine (Purinethol®)  thioguanine | PGx |
| *NUDT15* | azathioprine (Imuran®)  mercaptopurine (Purinethol®)  thioguanine | PGx |
| *DPYD* | capecitabine (Xeloda®)  fluorouracil (Adrucil®) | PGx |
| *UGT1A1* | atazanavir (Reyataz®)  belinostat (Beleodaq®)  Irinotecan (Camptosar®) | PGx |
| *SLCO1B1* | simvastatin (Zocor®) | PGx |
| *CYP2C19* | amitriptyline (Elaviil®)  citalopram (Celexa®)  clobazam (Onfi®)  clomipramine (Anafranil®)  clopidogrel (Plavix®)  doxepin (Sinequan®)  escitalopram (Lexapro®)  imipramine (Tofranil®)  setraline (Zoloft®)  trimipramine (Surmontil®)  voriconazole (Vfend®)  flibanserin (Addyi®)  pantoprazole (Protonix®)  brivaracetam (Briviact®) | PGx |
| *G6PD* | dabrafenib (Tafinlar®)  dapsone  hydroxychloroquine (Plaquenil®)  Local anesthetic containing drugs (e.g. articaine, chloroprocaine, lidocaine, mepivacaine, ropivacaine, tetracaine)  mafenide (Sulfamylon®)  methylene blue  nalidixic acid (NegGram®)  nitrofurantoin (Macrobid®, Macrodantin®, Furadentin®)  pegloticase (Krystexxa®)  phenazopyridine  primaquine  probenecid (Col-Benemid®)  rasburicase (Elitek®)  sodium nitrite  sulfacetamide  sulfamethoxazole/trimethoprim (Bactrim®, Septra®)  sulfanilamide  sulfasalazine (Azulfidine®)  tafenoquine (Krintafel®) | PGx |
| *APC* | Adenomatous polyposis coli (MIM 175100) | HDR |
| *MYH11* | Aortic aneurysm, familial thoracic 4 (MIM 132900) | HDR |
| *ACTA2* | Aortic aneurysm, familial thoracic 6 (MIM 611788) | HDR |
| *TMEM43* | Arrhythmogenic right ventricular cardiomyopathy, type 5 (MIM 604400) | HDR |
| *DSP* | Arrhythmogenic right ventricular cardiomyopathy, type 8 (MIM 607450) | HDR |
| *PKP2* | Arrhythmogenic right ventricular cardiomyopathy, type 9 (MIM 609040) | HDR |
| *DSG2* | Arrhythmogenic right ventricular cardiomyopathy, type 10 (MIM 610193) | HDR |
| *DSC2* | Arrhythmogenic right ventricular cardiomyopathy, type 11 (MIM 610476) | HDR |
| *BRCA1* | Breast-ovarian cancer, familial 1 (MIM 604370) | HDR |
| *BRCA2* | Breast-ovarian cancer, familial 2 (MIM 612555) | HDR |
| *SCN5A* | Brugada syndrome 1 (MIM 601144) | HDR |
| *RYR2* | Catecholaminergic polymorphic ventricular tachycardia (MIM 604772) | HDR |
| *LMNA* | Dilated cardiomyopathy 1A (MIM 115200) | HDR |
| *MYBPC3* | Dilated cardiomyopathy 1A (MIM 115200) | HDR |
| *COL3A1* | Ehlers-Danlos syndrome, type 4 (MIM 130050) | HDR |
| *GLA* | Fabry's disease (MIM 301500) | HDR |
| *APOB* | Familial hypercholesterolemia (MIM 143890) | HDR |
| *LDLR* | Familial hypercholesterolemia (MIM 143890) | HDR |
| *MYH7* | Familial hypertrophic cardiomyopathy 1 (MIM 192600) | HDR |
| *TPM1* | Familial hypertrophic cardiomyopathy 3 (MIM 115196) | HDR |
| *MYBPC3* | Familial hypertrophic cardiomyopathy 4 (MIM 115197) | HDR |
| *PRKAG2* | Familial hypertrophic cardiomyopathy 6 (MIM 600858) | HDR |
| *TNNI3* | Familial hypertrophic cardiomyopathy 7 (MIM 613690) | HDR |
| *MYL3* | Familial hypertrophic cardiomyopathy 8 (MIM 608751) | HDR |
| *MYL2* | Familial hypertrophic cardiomyopathy 10 (MIM 608758) | HDR |
| *ACTC1* | Familial hypertrophic cardiomyopathy 11 (MIM 612098) | HDR |
| *RET* | Familial medullary thyroid carcinoma (MIM 155240) | HDR |
| *PCSK9* | Hypercholesterolemia, autosomal dominant, 3 (MIM 603776) | HDR |
| *BMPR1A* | Juvenile polyposis syndrome, (MIM 174900) | HDR |
| *SMAD4* | Juvenile polyposis syndrome, (MIM 174900) | HDR |
| *TNNT2* | Left ventricular noncompaction 6 (MIM 601494) | HDR |
| *TP53* | Li-Fraumeni syndrome 1 (MIM 151623) | HDR |
| *TGFBR1* | Loeys-Dietz syndrome type 1A (MIM 609192) | HDR |
| *TGFBR2* | Loeys-Dietz syndrome type 1B (MIM 610168) | HDR |
| *TGFBR1* | Loeys-Dietz syndrome type 2A (MIM 608967) | HDR |
| *TGFBR2* | Loeys-Dietz syndrome type 2B (MIM 610380) | HDR |
| *SMAD3* | Loeys-Dietz syndrome type 3 (MIM 613795) | HDR |
| *KCNQ1* | Long QT syndrome 1 (MIM 192500) | HDR |
| *KCNH2* | Long QT syndrome 2 (MIM 613688) | HDR |
| *SCN5A* | Long QT syndrome 3 (MIM 603830) | HDR |
| *MLH1* | Lynch syndrome (MIM 120435) | HDR |
| *MSH2* | Lynch syndrome (MIM 120435) | HDR |
| *MSH6* | Lynch syndrome (MIM 120435) | HDR |
| *PMS2* | Lynch syndrome (MIM 120435) | HDR |
| *RYR1* | Malignant hyperthermia (MIM 145600) | HDR |
| *CACNA1S* | Malignant hyperthermia (MIM 145600) | HDR |
| *FBN1* | Marfan's syndrome (MIM 154700) | HDR |
| *TGFBR1* | Marfan's syndrome (MIM 154700) | HDR |
| *MEN1* | Multiple endocrine neoplasia, type 1 (MIM 131100) | HDR |
| *RET* | Multiple endocrine neoplasia, type 2a (MIM 171400) | HDR |
|  | Multiple endocrine neoplasia, type 2b (MIM 162300) | HDR |
| *MUTYH* | MYH-associated polyposis (MIM 608456) | HDR |
| *NF2* | Neurofibromatosis, type 2 (MIM 101000) | HDR |
| *OTC* | Ornithine carbamoyltransferase deficiency (MIM 311250) | HDR |
| *SDHD* | Paragangliomas 1 (MIM 168000) | HDR |
| *SDHAF2* | Paragangliomas 2 (MIM 601650) | HDR |
| *SDHC* | Paragangliomas 3 (MIM 605373) | HDR |
| *SDHB* | Paragangliomas 4 (MIM 115310) | HDR |
| *STK11* | Peutz-Jeghers syndrome (MIM 175200) | HDR |
| *MUTYH* | Pilomatrixoma (MIM 132600) | HDR |
| *PTEN* | PTEN hamartoma tumor syndrome (MIM 153480) | HDR |
| *RB1* | Retinoblastoma (MIM 180200) | HDR |
| *TSC1* | Tuberous sclerosis 1 (MIM 191100) | HDR |
| *TSC2* | Tuberous sclerosis 2 (MIM 613254) | HDR |
| *VHL* | Von Hippel-Lindau syndrome (MIM 193300) | HDR |
| *WT1* | Wilms' tumor (MIM 194070) | HDR |
| *ATP7B* | Wilson disease (MIM 277900) | HDR |
