## Supplemental Table 2 for "Whole genome sequencing as an investigational device for return of hereditary disease risk and pharmacogenomic results as part of the *All of Us* Research Program"

Table S2. Founder mutations in *BRCA1* and *BRCA2* called with 100% accuracy in independent samples

| **Sample ID** | **Gene** | **DNA change** | **Amino acid change** | **GRCh37 coordinates** | **Variant type** | **Zygosity** | **Site** |
| --- | --- | --- | --- | --- | --- | --- | --- |
| 1000070014 | *BRCA1* | c.68_69delAG | p.Glu23Valfs*17 | 17:41276045_41276046del | Deletion | Heterozygous | BCM*^a^* |
| 1000073979 | *BRCA1* | c.5266dupC | p.Gln1756Profs*74 | 17:41209080dupC | Insertion | Heterozygous | BCM |
| SM-IN9H2 | *BRCA1* | c.68_69delAG | p.Glu23Valfs*17 | 17:41276045_41276046delCT | Deletion | Heterozygous | BI*^b^* |
| SM-IN8QQ | *BRCA1* | c.5266dupC | p.Gln1756Profs*74 | 17:41209079_41209080insG | Insertion | Heterozygous | BI |
| SM-JPV8J | *BRCA1* | c.5266dupC | p.Gln1756Profs*74 | 17:41209079T>TG | Insertion | Heterozygous | BI |
| SM-JPV8Y | *BRCA1* | c.5266dupC | p.Gln1756Profs*74 | 17:41209079T>TG | Deletion | Heterozygous | BI |
| SM-JPVA4 | *BRCA1* | c.5266dupC | p.Gln1756Profs*74 | 17:41209079T>TG | Insertion | Heterozygous | BI |
| SM-JPVAE | *BRCA1* | c.5266dupC | p.Gln1756Profs*74 | 17:41209079T>TG | Insertion | Heterozygous | BI |
| 1000074456 | *BRCA2* | c.5946delT | p.Ser1982Argfs*22 | 13:32914438del | Deletion | Heterozygous | BCM |
| SM-GZQK6 | *BRCA2* | c.5946delT | p.Ser1982Argfs*22 | 13:32914438delT | Deletion | Heterozygous | BI |
| SM-JPTV9 | *BRCA2* | c.5946delT | p.Ser1982Argfs*22 | 13:32914437GT>G | Deletion | Heterozygous | BI |
| SM-JPV89 | *BRCA2* | c.5946delT | p.Ser1982Argfs*22 | 13:32914437GT>G | Deletion | Heterozygous | BI |
| SM-JPVA9 | *BRCA2* | c.5946delT | p.Ser1982Argfs*22 | 13:32914437GT>G | Deletion | Heterozygous | BI |
| 323249 | *BRCA2* | c.5946delT | p.Ser1982Argfs*22 | 13:32914438delT | Deletion | Heterozygous | UW*^c^* |

*^a^*BCM, Baylor College of Medicine. *^b^*BI, Broad Institute. *^c^*UW, University of Washington.
