## Supplemental Table 5 for "Whole genome sequencing as an investigational device for return of hereditary disease risk and pharmacogenomic results as part of the *All of Us* Research Program"

Table S5. PGx calling across additional cell line samples from the 1000 Genomes Project

| **Gene and star allele** | **PGx***^a^* **sample** | **Number of samples** | **Correct calls** | **Incorrect calls** | **Overall concordance** |
| --- | --- | --- | --- | --- | --- |
| *CYP2C19*16* | NA19452 | 1 | 1/1 | 0 | 1/1 |
| *CYP2C19*2* | HG02087 HG02511 NA18599 NA18617 NA18964 NA18978 NA18998 NA19000 NA19020 NA21110 | 10 | 10/10 | 0 | 10/10 |
| *CYP2C19*2/*2* | HG02186 HG02188 | 2 | 2/2 | 0 | 2/2 |
| *CYP2C19*22* | HG02318 | 1 | 1/1 | 0 | 1/1 |
| *CYP2C19*24* | NA20356 | 1 | 1/1 | 0 | 1/1 |
| *CYP2C19*3* | HG02141 HG02318 | 2 | 2/2 | 0 | 2/2 |
| *G6PD* Asahi | HG02511 | 1 | 1/1 | 0 | 1/1 |
| *G6PD* Aures | HG02165 | 1 | 1/1 | 0 | 1/1 |
| *G6PD* Canton, Taiwan-Hakka, Gifu-like, Agrigento-like | HG02087 NA18617 | 2 | 2/2 | 0 | 2/2 |
| *G6PD* Chinese-5 | HG02186 | 1 | 1/1 | 0 | 1/1 |
| *G6PD* Ilesha | NA19020 | 1 | 1/1 | 0 | 1/1 |
| *G6PD* Kalyan-Kerala, Jamnaga, Rohini | NA21110 | 1 | 1/1 | 0 | 1/1 |
| *G6PD* Seattle, Lodi, Modena, Ferrara-II, Athens-like | HG01620 | 1 | 1/1 | 0 | 1/1 |
| *G6PD* Sibari | NA19323 | 1 | 1/1 | 0 | 1/1 |
| *G6PD* Ube Konan | NA18998 | 1 | 1/1 | 0 | 1/1 |
| *G6PD* Viangchan, Jammu | HG02141 HG02188 | 2 | 2/2 | 0 | 2/2 |
| *NUDT15*2* | NA18599 NA18621 NA18622 NA18626 NA18633 NA18740 NA18964 NA18978 NA19000 NA19077 | 10 | 10/10 | 0 | 10/10 |
| *NUDT15*3* | NA18599 NA18998 | 2 | 2/2 | 0 | 2/2 |
| *SLCO1B1*15* | NA19000 | 1 | 1/1 | 0 | 1/1 |
| *SLCO1B1*17* | HG02186 HG02188 NA18599 NA19000 NA19077 | 5 | 5/5 | 0 | 5/5 |
| *TPMT*2* | HG01605 | 1 | 1/1 | 0 | 1/1 |
| *TPMT*3C* | NA18908 NA19114 NA19323 NA19452 | 4 | 4/4 | 0 | 4/4 |
| *UGT1A1*28* | NA18510 NA18908 NA19077 NA19323 | 4 | 4/4 | 0 | 4/4 |
| *UGT1A1*6* | HG02141 HG02188 NA18617 NA18621 NA18626 NA18633 NA18740 | 7 | 7/7 | 0 | 7/7 |

*^a^*PGx, pharmacogenomics.
