## Supplemental Table 6 for "Whole genome sequencing as an investigational device for return of hereditary disease risk and pharmacogenomic results as part of the *All of Us* Research Program"

Table S6. Overall equivalence of called variants in donor blood samples across the Genome Centers

|  |  | **SNV***^d^* **concordant** | **SNV discordant** | **InDel***^e^* **concordant** | **InDel discordant** | **Overall equivalence [95%CI***^f^***]** |
| --- | --- | --- | --- | --- | --- | --- |
| **Interlab** | **BCM***^a^***-UW***^b^* | 3830/3833 | 3/3833 | 189/195 | 6/195 | 99.77%  [99.55%-99.99%] |
|  | **BI***^c^***-BCM** | 3830/3834 | 4/3834 | 191/193 | 2/193 | 99.86%  [99.71%-100%] |
|  | **UW-BI** | 3831/3834 | 3/3834 | 188/196 | 8/196 | 99.71%  [99.46%-99.96%] |
| **Intralab** | **BI-BI** | 1708/1709 | 1/1709 | 87/89 | 2/89 | 99.82%  [99.57%-100%] |
|  | **BCM-BCM** | 1246/1248 | 2/1248 | 62/62 | 0/62 | 99.76%  [99.32%-100%] |
|  | **UW-UW** | 1577/1578 | 1/1578 | 76/83 | 7/83 | 99.49%  [99.02%-99.46%] |

*^a^*BCM, Baylor College of Medicine. *^b^*UW, University of Washington. *^c^*BI, Broad Institute. *^d^*SNV, single nucleotide variant. *^e^*InDel, insertion or deletion. *^f^*CI, Confidence interval.
