## Supplemental Table 7 for "Whole genome sequencing as an investigational device for return of hereditary disease risk and pharmacogenomic results as part of the *All of Us* Research Program"

Table S7. Overall equivalence of called variants in cell lines across the Genome Centers

|  |  | **SNV***^d^* **Concordant** | **SNV discordant** | **InDel***^e^* **concordant** | **InDel discordant** | **Overall equivalence [95%CI***^f^***]** |
| --- | --- | --- | --- | --- | --- | --- |
| **Interlab** | **BCM***^a^***-UW***^b^* | 24289/24304 | 15/24304 | 1142/1178 | 36/1178 | 99.80%  [99.71%-99.89%] |
|  | **BI***^c^***-BCM** | 24288/24304 | 16/24304 | 1151/1171 | 20/1171 | 99.86%  [99.79%-99.93%] |
|  | **UW-BI** | 24287/24302 | 15/24302 | 1145/1177 | 32/1177 | 99.81%  [99.73%-99.90%] |
| **Intralab** | **BI-BI** | 1175/1175 | 0/1175 | 56/65 | 9/65 | 99.27%  [99.68%-99.86%] |
|  | **BCM-BCM** | 806/806 | 0/806 | 38/40 | 2/40 | 99.76%  [99.32%-100%] |
|  | **UW-UW** | 918/918 | 0/918 | 50/50 | 0/50 | 100%  [100%-100%] |

*^a^*BCM, Baylor College of Medicine. *^b^*UW, University of Washington. *^c^*BI, Broad Institute. *^d^*SNV, single nucleotide variant. *^e^*InDel, insertion or deletion. *^f^*CI, confidence interval.
