## Supplemental Table 8 for "Whole genome sequencing as an investigational device for return of hereditary disease risk and pharmacogenomic results as part of the *All of Us* Research Program"

Table S8. Concordance of variant calling by genomic context in donor blood samples

| **Category** | **Genome Center** | **Panel+***^a^***/WGS–***^b^* | **Panel– /WGS+** | **Panel+ /WGS+** | **Panel– /WGS–** | **PPA***^c^* **[95% CI***^d^***]** | **NPA***^e^* **[95% CI]** |
| --- | --- | --- | --- | --- | --- | --- | --- |
| **SNVs***^f^* | BCM*^g^* | 7 | 13 | 2609 | 4187271 | 99.74% [99.6%-99.9%] | 100% [100%-100%] |
|  | BI*^h^* | 8 | 14 | 2610 | 4187268 | 99.71% [99.5%-99.9%] | 100% [100%-100%] |
|  | UW*^i^* | 8 | 13 | 2610 | 4187269 | 99.71% [99.5%-99.9%] | 100% [100%-100%] |
| **Insertions** | BCM | 1 | 20 | 20 | 4189859 | 97.22% [91.8%-100%] | 100% [100%-100%] |
|  | BI | 0 | 20 | 20 | 4189860 | 100% [100%-100%] | 100% [100%-100%] |
|  | UW | 2 | 20 | 20 | 4189858 | 94.44% [87.1%-100%] | 100% [100%-100%] |
| **Deletions** | BCM | 1 | 0 | 42 | 4189857 | 98.33% [95.1%-100%] | 100% [100%-100%] |
|  | BI | 1 | 0 | 42 | 4189857 | 98.33% [95.1%-100%] | 100% [100%-100%] |
|  | UW | 1 | 0 | 42 | 4189857 | 98.33% [95.1%-100%] | 100% [100%-100%] |
| **Segmental duplications** | BCM | 1 | 0 | 240 | 4189659 | 99.29% [97.9%-100%] | 100% [100%-100%] |
|  | BI | 1 | 0 | 240 | 4189659 | 99.29% [97.9%-100%] | 100% [100%-100%] |
|  | UW | 1 | 0 | 240 | 4189659 | 99.29% [97.9%-100%] | 100% [100%-100%] |
| **Low mappability regions** | BCM | 1 | 0 | 24 | 3770885 | 97.22% [91.8%-100%] | 100% [100%-100%] |
|  | BI | 1 | 0 | 24 | 3770885 | 97.22% [91.8%-100%] | 100% [100%-100%] |
|  | UW | 1 | 0 | 24 | 3770885 | 97.22% [91.8%-100%] | 100% [100%-100%] |
| **Low complexity**  **regions** | BCM | 0 | 0 | 36 | 4189864 | 100% [100%-100%] | 100% [100%-100%] |
|  | BI | 0 | 0 | 36 | 4189864 | 100% [100%-100%] | 100% [100%-100%] |
|  | UW | 0 | 0 | 36 | 4189864 | 100% [100%-100%] | 100% [100%-100%] |
| **Low GC regions** | BCM | 0 | 0 | 20 | 1675940 | 100% [100%-100%] | 100% [100%-100%] |
|  | BI | 0 | 0 | 20 | 1675940 | 100% [100%-100%] | 100% [100%-100%] |
|  | UW | 0 | 0 | 20 | 1675940 | 100% [100%-100%] | 100% [100%-100%] |
| **Heterozygous variants** | BCM | 11 | 15 | 1648 | 4188226 | 99.37% [99.0%-99.7%] | 100% [100%-100%] |
|  | BI | 13 | 16 | 1648 | 4188223 | 99.27% [98.9%-99.7%] | 100% [100%-100%] |
|  | UW | 13 | 15 | 1648 | 4188224 | 99.27% [98.9%-99.7%] | 100% [100%-100%] |
| **Homozygous variants** | BCM | 1 | 18 | 1025 | 4188856 | 99.92% [99.8%-100%] | 100% [100%-100%] |
|  | BI | 0 | 18 | 1026 | 4188856 | 100% [100%-100%] | 100% [100%-100%] |
|  | UW | 0 | 18 | 1026 | 4188856 | 100% [100%-100%] | 100% [100%-100%] |

*^a^+* indicates that a variant was present on the panel or the corresponding whole genome sequencing (WGS) sample. *^b^*– indicates that a variant was not present on the panel or the corresponding WGS sample. *^c^*PPA, positive percent agreement. *^d^*CI, confidence interval. *^e^*NPA, negative percent agreement. *^f^*SNVs, single nucleotide variants. *^g^*BCM, Baylor College of Medicine. *^h^*BI, Broad Institute. *^i^*UW, University of Washington.
