## Supplemental Table 9 for "Whole genome sequencing as an investigational device for return of hereditary disease risk and pharmacogenomic results as part of the *All of Us* Research Program"

Table S9. Concordance of variant calling by genomic context in cell lines

| **Category** | **Genome Center** | **Panel+***^a^***/WGS–***^b^* | **Panel– /WGS+** | **Panel+ /WGS+** | **Panel– /WGS–** | **PPA***^c^* **[95% CI***^d^***]** | **NPA***^e^* **[95% CI]** |
| --- | --- | --- | --- | --- | --- | --- | --- |
| **SNVs***^f^* | BCM*^g^* | 24 | 1 | 3558 | 6071772 | 99.34% [99.0%-99.7%] | 100% [100%-100%] |
|  | BI*^h^* | 24 | 1 | 3694 | 6281131 | 99.36% [99.1%-99.7%] | 100% [100%-100%] |
|  | UW*^i^* | 24 | 0 | 3694 | 6281132 | 99.36% [99.1%-99.7%] | 100% [100%-100%] |
| **Insertions** | BCM | 1 | 23 | 21 | 5237330 | 96.70% [90.1%-100%] | 100% [100%-100%] |
|  | BI | 1 | 22 | 22 | 5446825 | 96.88% [90.8%-100%] | 100% [100%-100%] |
|  | UW | 1 | 23 | 22 | 5446824 | 96.88% [90.8%-100%] | 100% [100%-100%] |
| **Deletions** | BCM | 1 | 9 | 63 | 5865787 | 99.11% [97.4%-100%] | 100% [100%-100%] |
|  | BI | 1 | 9 | 65 | 6075280 | 99.14% [97.4%-100%] | 100% [100%-100%] |
|  | UW | 1 | 10 | 65 | 6075279 | 99.14% [97.4%-100%] | 100% [100%-100%] |
| **Segmental duplications** | BCM | 0 | 0 | 309 | 6075046 | 100% [100%-100%] | 100% [100%-100%] |
|  | BI | 0 | 1 | 322 | 6284527 | 100% [100%-100%] | 100% [100%-100%] |
|  | UW | 0 | 0 | 322 | 6284528 | 100% [100%-100%] | 100% [100%-100%] |
| **Low mappability regions** | BCM | 0 | 0 | 16 | 2932915 | 100% [100%-100%] | 100% [100%-100%] |
|  | BI | 0 | 0 | 18 | 3142407 | 100% [100%-100%] | 100% [100%-100%] |
|  | UW | 0 | 0 | 18 | 3142407 | 100% [100%-100%] | 100% [100%-100%] |
| **Low complexity regions** | BCM | 0 | 11 | 54 | 6075290 | 100% [100%-100%] | 100% [100%-100%] |
|  | BI | 0 | 11 | 54 | 6075290 | 100% [100%-100%] | 100% [100%-100%] |
|  | UW | 0 | 10 | 54 | 6075291 | 100% [100%-100%] | 100% [100%-100%] |
| **Low GC regions** | BCM | 1 | 0 | 34 | 4399360 | 99.21% [97.7%-100%] | 100% [100%-100%] |
|  | BI | 1 | 0 | 36 | 4608853 | 99.24% [97.8%-100%] | 100% [100%-100%] |
|  | UW | 1 | 0 | 36 | 4608853 | 99.24% [97.8%-100%] | 100% [100%-100%] |
| **Heterozygous variants** | BCM | 18 | 20 | 2225 | 6073092 | 99.22% [98.7%-99.7%] | 100% [100%-100%] |
|  | BI | 18 | 20 | 2323 | 6282489 | 99.24% [98.8%-99.7%] | 100% [100%-100%] |
|  | UW | 18 | 20 | 2323 | 6282489 | 99.24% [98.8%-99.7%] | 100% [100%-100%] |
| **Homozygous variants** | BCM | 8 | 13 | 1417 | 6073917 | 99.52% [99.1%-100%] | 100% [100%-100%] |
|  | BI | 8 | 12 | 1458 | 6283372 | 99.54% [99.1%-100%] | 100% [100%-100%] |
|  | UW | 8 | 13 | 1458 | 6283371 | 99.54% [99.1%-100%] | 100% [100%-100%] |

*^a^+* indicates that a variant was present on the panel or the corresponding whole genome sequencing (WGS) sample. *^b^*– indicates that a variant was not present on the panel or the corresponding WGS sample. *^c^*PPA, positive percent agreement. *^d^*CI, confidence interval. *^e^*NPA, negative percent agreement. *^f^*SNVs, single nucleotide variants. *^g^*BCM, Baylor College of Medicine. *^h^*BI, Broad Institute. *^i^*UW, University of Washington.
