## Supplemental Table 10 for "Whole genome sequencing as an investigational device for return of hereditary disease risk and pharmacogenomic results as part of the *All of Us* Research Program"

Table S10. Equivalence of performance measures between cell line-derived DNA and blood-derived DNA

| **Performance measures** | | | | | | | | | |
| --- | --- | --- | --- | --- | --- | --- | --- | --- | --- |
|  | **Accuracy (n)** | **P/LP***^a^* **variant accuracy** | **PGx***^b^* **allele accuracy** | **Precision (n)** | **Interlab concordance (BCM***^c^***-UW***^d^***) [95% CI***^e^***]** | | **Interlab concordance (BI***^f^***-BCM) [95% CI]** | | **Interlab concordance (UW-BI) [95% CI]** |
| **Clinical samples** | 271 | 100% | 100% | 28 | 99.77%  [99.55%-99.99%] | | 99.86%  [99.71%-100%] | | 99.71%  [99.46%-99.96%] |
| **Cell line samples** | 30 | 100% | 100% | 175 | 99.8%  [99.71%-99.89%] | | 99.86%  [99.79%-99.93%] | | 99.81%  [99.73%-99.90%] |

*^a^*Pathogenic/likely pathogenic. *^b^*PGx, pharmacogenomics. *^c^*BCM, Baylor College of Medicine. *^d^*UW, University of Washington. *^f^*CI, confidence interval. *^f^*BI, Broad Institute.
