## Supplemental Table 11 for "Whole genome sequencing as an investigational device for return of hereditary disease risk and pharmacogenomic results as part of the *All of Us* Research Program"

Table S11. Equivalence technical metrics between cell line-derived DNA and blood-derived DNA

| **Technical metrics** | | | | | | | | | | |
| --- | --- | --- | --- | --- | --- | --- | --- | --- | --- | --- |
|  | **Number of samples** | **% aligned bases**  **[std dev***^a^***]** | **% duplicate reads**  **[std dev]** | **Insert size (bp)**  **[std dev]** | **% Q30 bases**  **[std dev]** | **% chimeric reads**  **[std dev]** | **Genome coverage [std dev]** | **% covered ≥20X**  **[std dev]** | **% contam- ination**  **[std dev]** | **Ti/Tv ratio**  **[std dev]** |
| **Clinical samples** | 253 | 92%  [±0.94%] | 11.15%  [±1.78%] | 421 bp  [±16 bp] | 92.2%  [±0.77%] | 1.81%  [±0.24%] | 39.99X  [±7.3X] | 96.55%  [±0.49%] | 0.03%  [±0.33%] | 1.94  [±0.01] |
| **Cell line samples** | 223 | 91.25%  [±0.93%] | 11.14%  [±1.4%] | 419 bp  [±11 bp] | 91.43%  [±1.03%] | 1.69%  [±0.16%] | 39.53X  [±3.13X] | 96.07%  [±0.24%] | 0.02%  [±0.07%] | 1.93  [± 0.01] |

*^a^*Std dev, standard deviation.
