## Supplemental Table 13 for "Whole genome sequencing as an investigational device for return of hereditary disease risk and pharmacogenomic results as part of the *All of Us* Research Program"

Table S13. Input titration results for four blood donor samples

| **Input (ng)** | **Library construction success rate** | **Mean sensitivity** | **Std dev***^a^* **sensitivity** | **Mean precision** | **Std dev precision** |
| --- | --- | --- | --- | --- | --- |
| 25 | 0% | n/a^b^ | n/a | n/a | n/a |
| 100 | 17% | 99% | n/a | 100% | n/a |
| 250 | 100% | 100% | 0% | 100% | 0% |
| 375 | 100% | 98% | 1% | 99% | 0% |
| 500 | 100% | 99% | 1% | 99% | 1% |
| 750 | 100% | 98% | 1% | 99% | 0% |
| 1500 | 100% | 98% | 1% | 98% | 0% |

*^a^*Std dev, standard deviation.^b^n/a = not applicable due to no data or not enough data.
