## Supplemental Table 14 for "Whole genome sequencing as an investigational device for return of hereditary disease risk and pharmacogenomic results as part of the *All of Us* Research Program"

Table S14. Frequently underperforming bases within the Hereditary Disease Risk Report

| **Gene** | **Total sites** | **GRCh37**  **low-coverage sites** | **GRCh38**  **low-coverage sites** |
| --- | --- | --- | --- |
| *MYH11* | 6699 | 8 | 8 |
| *MSH2* | 3148 | 14 | 14 |
| *KCNH2* | 3131 | 24 | 23 |
| *TSC1* | 3936 | 10 | 10 |
