## Supplemental Table 15 for "Whole genome sequencing as an investigational device for return of hereditary disease risk and pharmacogenomic results as part of the *All of Us* Research Program"

Table S15. Fail rates for samples at discrete parts of the process across the Genome Centers

| **Genome Centers (WGS***^a^***)** | | | | | | | | | | | | |
| --- | --- | --- | --- | --- | --- | --- | --- | --- | --- | --- | --- | --- |
| **Lab** | **Cohort** | **Instrument** | **Sample intake** | | **Library** | | **Sequencing** | | **Data quality** | | **Total samples failed** | **Aggregate sample fail rate** |
|  |  |  | **Input #** | **# failed (% of total fails)** | **Input #** | **# failed (% of total fails)** | **Input #** | **# failed (% of total fails)** | **Input #** | **# failed (% of total fails)** |  |  |
| **BCM***^b^* | TOPMed*^f^* | NovaSeq | 1986 | 103 (90.4%) | 1883 | 0 (0%) | 1883 | 0 (0%) | 1883 | 11 (9.6%)*^e^* | 114 | 5.74% |
|  | AoURP*^g^* | NovaSeq | 584 | 0 (0%) | 584 | 0 (0%) | 584 | 0 (0%) | 584 | 1 (100%) | 1 | 0.17% |
| **Broad***^c^* | TOPMed | NovaSeq | 6391 | 547 (99.5%) | 5844 | 3 (0.5%) | 5841 | 0 (0%) | 5841 | 0 (0%) | 550 | 8.61% |
|  | AoURP | NovaSeq | 424 | 3 (33.3%) | 421 | 0 (0%) | 421 | 0 (0%) | 421 | 6 (66.7%) | 9 | 2.12% |
| **UW***^d^* | TOPMed | HiSeqX | 14587 | 1346 (88.3%) | 13241 | 45 (3%) | 13196 | 102 (6.7%) | 13094 | 32 (2%) | 1,525 | 10.45% |
|  | AoURP | NovaSeq | 291 | 0 (0%) | 291 | 0 (0%) | 291 | 2 (33.3%) | 289 | 4 (66.7%) | 6 | 2.06% |

*^a^*WGS, whole genome sequencing. *^b^*Baylor College of Medicine. *^c^*Broad Institute. *^d^*University of Washington. *^e^*BCM sample fails are in amplicon generation. *^f^*National Heart, Lung, and Blood Institute Trans-Omics for Precision Medicine. *^g^All of Us* Research Program.
