## Supplemental Table 16 for "Whole genome sequencing as an investigational device for return of hereditary disease risk and pharmacogenomic results as part of the *All of Us* Research Program"

Table S16. Fail rates for samples at discrete parts of the process across the Clinical Validation Laboratories

| **CVLs***^a^* | | | | | | | | | | | | |
| --- | --- | --- | --- | --- | --- | --- | --- | --- | --- | --- | --- | --- |
| **Lab** | **Cohort** | **Instrument** | **Sample Intake** | | **Library** | | **Sequencing** | | **Data Quality** | | **Total samples failed** | **Aggregate sample fail rate** |
|  |  |  | **Input #** | **# Failed (% of total fails)** | **Input #** | **# Failed (% of total fails)** | **Input #** | **# failed (% of total fails)** | **Input #** | **# failed (% of total fails)** |  |  |
| **BCM***^b^* | Internal | ABI 3730/3500 | 1635 | 0 (0%) | NA | NA | 1635 | 0 (0%) | 1635 | 0 (0%) | 10*^d^* | 0.61% |
| **Color** | AoURP*^d^* | NovaSeq | 114 | 0 (0%) | 114 | 1 (100%) | 113 | 0 (0%) | 113 | 0 (0%) | 1 | 0.88% |
| **UW***^c^* | AoURP | NovaSeq | 315 | 0 (0%) | 315 | 0 (0%) | 315 | 2 (50%) | 313 | 2 (50%) | 4 | 1.27% |

*^a^*CVLs, Clinical Validation Laboratories.*^b^*BCM, Baylor College of Medicine. *^c^*UW, University of Washington. *^d^All of Us* Research Program.
