## Supplemental Table 17 for "Whole genome sequencing as an investigational device for return of hereditary disease risk and pharmacogenomic results as part of the *All of Us* Research Program"

Table S17. Feedback post IDE submission

| **Feedback area** | **Post-submission United States Food and Drug Administration feedback** |
| --- | --- |
| Program organization | Please provide a description of the enrollment, informed consent, and biospecimen collection procedures. |
|  | Clarify whether all Clinical Validation Laboratories have the same roles. |
|  | Please describe any processes/procedures in place to ensure that bias is not introduced during the resolution of variant harmonization disagreements. |
|  | Please provide the informing loop information that will be provided to subjects for our review. |
|  | Please provide a detailed description of your procedures/processes for updating variant classifications. |
|  | Describe how data will be delivered to participants, including details about the participant portal interface. |
|  | Please provide all educational materials that will be provided/available to subjects for our review. |
|  | Please clarify in which situations the genetic counselling resource will provide support and the level of support that will be provided, particularly in the case of a significant re-classification in either direction. |
| Report design / language | You should include in your reports that there is a possibility that the variant interpretation of the test result provided by your investigational device is incorrect. |
|  | Do not include any language that may imply to a subject that the research result from your investigational device is actionable. |
|  | Provide the report language that will be used to describe a positive finding in each reportable gene. |
| Study design | The United States Food and Drug Administration considers the study investigational in nature (i.e., results from your study are not generated from a cleared or approved assay validated for clinical use). Revise your protocol to consistently convey that your study is an investigational device study. |
|  | All risks of the study should be conveyed together in the consent. |
|  | Please clarify the reporting process for subjects who are Hereditary Disease Risk positive for multiple genes. |
|  | Clarify what raw data types the participants may elect to receive. |
|  | For pharmacogenomic gene-drug associations, do not report associations with ‘moderate’ evidence. |
| Technical information | Please provide a description of parameter settings for running the bioinformatics pipeline and ensure that each sequencing center is using the same parameter. These parameters should be locked throughout the investigational device study |
| Validation study requirements | Provide the false negative rate for the device |
|  | Please provide a detailed description of the orthogonal method used to generate ‘ground truth’ in the accuracy study and your justification for why this assay was selected. |
|  | Justify the number of control samples used for *G6PD* Asahi. |
|  | Clarify/explain a specific discordance between the *UGT1A1**28/*28 ground truth and validation data. |
|  | Please clarify if you have evaluated the reproducibility of your investigational device, in terms of reagent lot to lot, instrument to instrument, operator to operator, and day to day reproducibility. |
|  | Please provide protocols or detailed descriptions of the experimental designs and data analysis methods sufficient to understand how the studies were conducted. |
|  | You should include information in your investigational device exemption submission to demonstrate that the alleles you have chosen are adequate to represent the alleles your investigational device is intended to detect. |
|  | If you have a protocol (with acceptance criteria) describing the validation of new genes to be added to your investigational device in the future, please provide it |
|  | If you have any information about substances known to interfere with your device technology and intended sample type, please provide it. |
|  | Please clarify the patient sample types (anticoagulated whole blood, saliva, etc.) used for this study. |
|  | Please provide a breakdown of the user comprehension study results for each survey question by education level (e.g., breakdown of education levels for those who answered incorrectly for each survey question). |
|  | Please describe any discordant results from these precision studies. |
