## Supplemental Table 18 for "Whole genome sequencing as an investigational device for return of hereditary disease risk and pharmacogenomic results as part of the *All of Us* Research Program"

Table S18. Excluded recurrent false positive and false negative variants

| **Gene** | **Variant start site** |
| --- | --- |
| *MSH2* | chr2:47641559 |
| *MSH2* | chr2:47641562 |
| *APOB* | chr2:21266774 |
| *PMS2* | chr7:6037057 |
| *TSC1* | chr9:135773000 |
| *PCSK9* | chr1:55505552 |
