## Supplemental Table 19 for "Whole genome sequencing as an investigational device for return of hereditary disease risk and pharmacogenomic results as part of the *All of Us* Research Program"

Table S19. Accuracy of variant calls across the reportable region of NA12878

| **Reportable region** | **TP***^a^* | **TN***^b^* | **FP***^c^* | **FN***^d^* | **PPV***^e^* **[95% CI***^f^***]** | **NPV***^g^* **[95% CI]** |
| --- | --- | --- | --- | --- | --- | --- |
| All variants | 131 | 218551 | 0 | 0 | 100% [97.2% - 100%] | 100% [100% - 100%] |
| SNVs*^h^* only | 129 | 218551 | 0 | 0 | 100% [97.2% - 100%] | 100% [100% - 100%] |
| Insertions and deletions | 2 | 218551 | 0 | 0 | *** | 100% [100% - 100%] |
| Segmental duplications | 12 | 19073 | 0 | 0 | 100% [73.5% - 100%] | 100% [100% - 100%] |
| Low mappability regions | 1 | 3532 | 0 | 0 | *** | 100% [100% - 100%] |
| Low complexity regions | 4 | 2435 | 0 | 0 | *** | 100% [100% - 100%] |
| Low GC regions | 2 | 2722 | 0 | 0 | *** | 100% [100% - 100%] |
| High GC regions | 0 | 7 | 0 | 0 | *** | 100% [100% - 100%] |
| Heterozygous variants only | 82 | 218551 | 0 | 0 | 100% [95.6% - 100%] | 100% [100% - 100%] |
| Homozygous variants only | 49 | 218551 | 0 | 0 | 100% [92.7% -100%] | 100% [100% - 100%] |

*^a^*TP, true positive. *^b^*TN, true negative. *^c^*FP, false positive. *^d^*FN, false negative. *^e^*PPV, positive predictive value. *^f^*CI, confidence interval. *^g^*NPV, negative predictive agreement. *^h^*SNVs, single-nucleotide variants.
