## Supplemental Table 20 for "Whole genome sequencing as an investigational device for return of hereditary disease risk and pharmacogenomic results as part of the *All of Us* Research Program"

Table S20. Accuracy of variant calls across the whole genome of NA12878

| **Whole genome** | **TP***^a^* | **TN***^b^* | **FP***^c^* | **FN***^d^* | **PPV***^e^* **[95% CI***^f^***]** | **NPV***^g^* **[95% CI]** |
| --- | --- | --- | --- | --- | --- | --- |
| All variants | 3688413 | 2571369697 | 3907 | 2448 | 99.89% [99.89% - 99.90%] | 100% [100% - 100%] |
| SNVs*^h^* only | 3207969 | 2571369697 | 2522 | 1346 | 99.92% [99.92% - 99.92%] | 100% [100% - 100%] |
| Insertions and deletions | 480444 | 2571369697 | 1385 | 1102 | 99.71% [99.70% - 99.73%] | 100% [100% - 100%] |
| Segmental duplications | 59054 | 43102836 | 375 | 329 | 99.37% [99.30% - 99.43%] | 100% [100% - 100%] |
| Low mappability regions | 172097 | 99469838 | 1458 | 1296 | 99.16% [99.12% - 99.20%] | 100% [100% - 100%] |
| Low complexity regions | 257660 | 62578838 | 879 | 823 | 99.66% [99.64% - 99.68%] | 100% [100% - 100%] |
| Low GC regions | 179834 | 120814998 | 276 | 211 | 99.85% [99.83% - 99.86%] | 100% [100% - 100%] |
| High GC regions | 413 | 377579 | 5 | 0 | 98.80% [97.23% - 99.61%] | 100% [100% - 100%] |
| Heterozygous variants only | 2233008 | 2571369697 | 3614 | 1621 | 99.84% [99.83% - 99.84%] | 100% [100% - 100%] |
| Homozygous variants only | 1454259 | 2571369697 | 256 | 790 | 99.98% [99.98% - 99.98%] | 100% [100% - 100%] |

*^a^*TP, true positive. *^b^*TN, true negative. *^c^*FP, false positive. *^d^*FN, false negative. *^e^*PPV, positive predictive value. *^f^*CI, confidence interval. *^g^*NPV, negative predictive agreement. *^g^*SNVs, single-nucleotide variants.
