## Supplemental Table 21 for "Whole genome sequencing as an investigational device for return of hereditary disease risk and pharmacogenomic results as part of the *All of Us* Research Program"

Table S21. Performance of different extraction platforms and input material types

| **Category** | **Type** | **Panel+***^a^***/WGS–***^b^* | **Panel– /WGS+** | **Panel+ /WGS+** | **Panel– /WGS–** | **PPA***^c^* **[95% CI***^d^***]** | **NPA***^e^* **[95% CI]** |
| --- | --- | --- | --- | --- | --- | --- | --- |
| Autogen | SNV*^g^* | 11 | 29 | 4340 | 3138045 | 99.8% [99.6%-99.9%] | 100% [100%-100%] |
| Chemagen | SNV | 9 | 11 | 3489 | 3138916 | 99.8% [99.6%-99.9%] | 100% [100%-100%] |
| Autogen | InDel*^h^* | 3 | 33 | 114 | 3142275 | 98.1% [96.1%-100%] | 100% [100%-100%] |
| Chemagen | InDel | 0 | 26 | 72 | 3142327 | 100% [100%-100%] | 100% [100%-100%] |
| WBC*^f^* | SNV | 11 | 11 | 3816 | 3138587 | 99.7% [99.5%-99.8%] | 100% [100%-100%] |
| Whole Blood | SNV | 12 | 29 | 4013 | 3138371 | 99.7% [99.5%-99.9%] | 100% [100%-100%] |
| WBC | InDel | 3 | 29 | 90 | 3142303 | 98.5% [96.8%-100%] | 100% [100%-100%] |
| Whole Blood | InDel | 0 | 30 | 96 | 3142299 | 100% [100%-100%] | 100% [100%-100%] |

*^a^+* indicates that a variant was present on the panel or the corresponding whole genome sequencing (WGS) sample. *^b^*– indicates that a variant was not present on the panel or the corresponding WGS sample. *^c^*PPA, positive percent agreement. *^d^*CI, confidence interval. *^e^*NPA, negative percent agreement. *^f^*WBC, whole blood and buffy coat. *^g^*SNVs, single nucleotide variants. *^h^*InDel, insertion or deletion.
