## Supplemental Table 22 for "Whole genome sequencing as an investigational device for return of hereditary disease risk and pharmacogenomic results as part of the *All of Us* Research Program"

Table S22. Mismatched bases after liftover between reference builds

|  | **GRCh37** | | | | **GRCh38** | | | |
| --- | --- | --- | --- | --- | --- | --- | --- | --- |
| **Gene** | **Chr** | **Start** | **End** | **Sequence** | **Chr** | **Start** | **End** | **Sequence** |
| *APOB* | chr2 | 21,235,474 | 21,235,475 | T | chr2 | 21,012,602 | 21,012,603 | C |
| *DSP* | chr6 | 7,563,982 | 7,563,983 | T | chr6 | 7,563,749 | 7,563,750 | G |
| *FBN1* | chr15 | 48,807,636 | 48,807,637 | C | chr15 | 48,515,439 | 48,515,440 | T |
| *TNNI3* | chr19 | 55,665,583 | 55,665,584 | A | chr19 | 55,154,215 | 55,154,216 | C |
