## Supplemental Table 23 for "Whole genome sequencing as an investigational device for return of hereditary disease risk and pharmacogenomic results as part of the *All of Us* Research Program"

Table S23. Change management process, by risk categorization

| **Category** | **Possible types of changes** | **Definition / Examples** | **Change level** | **Type of FDA notification** |
| --- | --- | --- | --- | --- |
| **Enrollment** | Significant clinical protocol change | Change in intended use / indication, reduction in sample size, change in method of estimation, early termination of study | Major | Supplement |
|  | Consent form content | Change of consent as new risk of results is identified, collection procedure | Moderate | 5-day notice |
| **Specimen collection** | Specimen collection kits | New brand of collection kits (if switching from a validated kit to a different product validated for the same use) | Minor | Annual |
| **Specimen quality** | New acceptable specimen type | Adding saliva or buccal swabs | Major | Supplement |
|  | Specimen intake rejection criteria | The Biobank adding a policy to reject frozen blood samples | Major | Supplement |
|  | Specimen intake procedures | The Biobank changing shipping instructions, courier | Minor | Annual |
| **DNA quality and quantity** | Acceptance criteria | Adjust the cut-off of A260/280, quantity or quality of DNA | Major | Supplement |
|  | Quantification method | Using a new method other than Picogreen | Minor | Annual |
| **Instruments and methods** | Sequencer change | Using sequencing other than Novaseq | Major | Supplement |
|  | Instrument script/program changes | Scripts change to optimize liquid handling | Minor | Annual |
| **Pipeline** | Components of pipeline | Changing the variant caller, aligner, reference genome | Moderate | 5-day notice |
|  | Reportable variant types | Adding structural or copy number variants | Major | Supplement |
|  | Software version updates | Bioinformatic pipeline updates | Minor | Annual |
| **Report** | Cosmetic format changes | *All of Us* logo revisions, moving sections within the report, adding a section which does not include clinical information | Minor | Annual |
|  | Return of results process | The Genome Centers return reports directly to participants | Major | Supplement |
