## Supplemental Table 24 for "Whole genome sequencing as an investigational device for return of hereditary disease risk and pharmacogenomic results as part of the *All of Us* Research Program"

Table S24. Lessons learned during the pre-submission process

| **Initial expectation** | **Corrected Expectations / Components of final IDE application** |
| --- | --- |
| Clearly identifying results as part of a research study mitigates clinical risk. | The research nature of the program does mitigate risk, but throughout the IDE process, we had to make significant specific changes to the text to emphasize this point. |
| Pre-analytical (e.g. biobanking) and post-analytical steps (e.g., confirmatory testing, report generation) are not a part of a WGS*^a^* ‘medical device’. | The entire genomic workflow from sample receipt to return of results was included in the IDE |
| The program would be composed of multiple investigational devices. | The program would be composed of a single investigational device. To that end, the Genome Centers performed several studies to demonstrate precision across sites. |
| Blood and saliva samples would be included in the submission. | Only blood samples were included as additional studies were required for saliva as a sample type. |
| Control samples could be used to determine sensitivity, specificity, positive predictive value, repeatability, reproducibility, and limit of detection. | Validation criteria must be broken down by variant type and genomic context. |
| Samples used in validation studies should provide coverage of typical reporting results. | Validation cohort must reflect the variant prevalence of the population being tested. 'Highly prevalent' variants should make up most of the validation set. The set of highly prevalent, clinically significant variants should be pre-specified. |
| Control samples from cell lines are acceptable. | Most data should come from samples of the same type that will be collected by the program. |
| Standards and guidelines from expert bodies relating to pharmacogenomics reporting could be used in the selection of reportable alleles. | Information conveyed to participants should be limited to drugs for which there is information in the FDA*^b^* approved drug labeling that describes how genetic information can be used. |
| Standard clinical genetic testing reporting practices are acceptable. | The program must demonstrate that participants understand the reports and perform comprehension testing in a genetic testing-naive cohort that reflects the diversity of the program’s participants. |
| Future validations out of scope of the IDE.*^c^* | An explanation of how reporting will change in the future was included. |
| Existing quality metrics used for clinical laboratories were acceptable. | For quality metrics, a data-driven explanation of the specific cutoff selection was required. |
| There is a set standard to meet, as determined by the FDA. | With no predicate test, developing the standard to meet was a highly iterative process in collaboration with the FDA. |

*^a^*WGS, whole genome sequencing. *^b^*FDA, United States Food and Drug Administration. *^c^*IDE, investigational device exemption.
